# Estimating epidemiological quantities from repeated cross-sectional prevalence measurements

**DOI:** 10.1101/2022.03.29.22273101

**Authors:** Sam Abbott, Sebastian Funk

## Abstract

**Background:** Repeated measurements of cross-sectional prevalence of Polymerase Chain Reaction (PCR) positivity or seropositivity provide rich insight into the dynamics of an infection. The UK Office for National Statistics (ONS) Community Infection Survey publishes such measurements for SARS-CoV-2 on a weekly basis based on testing enrolled households, contributing to situational awareness in the country. Here we present estimates of time-varying and static epidemiological quantities that were derived from the estimates published by ONS.

**Methods:** We used a gaussian process to model incidence of infections and then estimated observed PCR prevalence by convolving our modelled incidence estimates with a previously published PCR detection curve describing the probability of a positive test as a function of the time since infection. We refined our incidence estimates using time-varying estimates of antibody prevalence combined with a model of antibody positivity and waning that moved individuals between compartments with or without antibodies based on estimates of new infections, vaccination, probability of seroconversion and waning.

**Results:** We produced incidence curves of infection describing the UK epidemic from late April 2020 until early 2022. We used these estimates of incidence to estimate the time-varying growth rate of infections, and combined them with estimates of the generation interval to estimate time-varying reproduction numbers. Biological parameters describing seroconversion and waning, while based on a simple model, were broadly in line with plausible ranges from individual-level studies.

**Conclusions:** Beyond informing situational awareness and allowing for estimates using individual-level data, repeated cross-sectional studies make it possible to estimate epidemiological parameters from population-level models. Studies or public health surveillance methods based on similar designs offer opportunities for further improving our understanding of the dynamics of SARS-CoV-2 or other pathogens and their interaction with population-level immunity.

## Introduction

Infectious disease surveillance serves to monitor the health of populations and identify new threats as quickly as possible after they arise (Murray & Cohen, 2017). It is often based on healthcare-based reporting systems whereby primary care providers or hospitals report numbers of individuals identified as likely cases of a disease to central authorities where these numbers are collated and reported as aggregates. During the Covid-19 pandemic in the United Kingdom, reporting of cases has mostly involved collating numbers of laboratory-identified infections with SARS-CoV-2 via self-reporting, community testing sites or hospitals.

A separate and independent system of collating information on the state of the pandemic has been run by the Office for National Statistics (ONS) via its Community Infection Survey, which conducts repeated cross-sectional surveys of Polymerase Chain Reaction (PCR) positivity indicating infection with SARS-CoV-2, as well as antibody seroprevalence via household visits (Pouwels et al., 2020). By adjusting for biases in the sampled population, the study has been used to estimate daily population-wide estimates of infection prevalence, unaffected by testing capacity or reporting behaviour that often varies by age as well as sociodemographic or other factors.

While repeated randomised cross-sectional sampling of positivity and antibodies provides utility in themselves for tracking an epidemic in real time, they can also be used for estimating epidemiological quantities by combining them with information on infection kinetics and immunological responses. Here we present a semi-mechanistic model that combines PCR positivity curves, generation interval estimates and vaccination data with ONS PCR positivity and antibody data to estimate infection incidence and its growth rates, reproduction numbers and rates of antibody waning.

## Methods

### Data

We obtained the published estimates of daily prevalence of Polymerase Chain Reaction (PCR) positivity beginning on 26 April, 2020, from the ONS Community infection survey separately by nation, region, age group and variant, alongside their 95% credible intervals, from the published spreadsheets on the ONS web site. ONS estimates of a given prevalence vary between publication dates as the internal model to calculate prevalence involves smoothing, such that new data points in the present affect the estimates of times past. We aggregated estimates of PCR positivity for a single day produced for different publication dates by calculating the central estimate and credible intervals as the medians of the different respective central estimates and credible intervals.

### Model

We developed a Bayesian model to estimate epidemiological quantities from ONS PCR positivity estimates and, optionally, population level antibody prevalence estimates and vaccination coverage.

#### PCR positivity

We estimated the population proportion newly infected in the population *I*(*t*) as a latent variable that is convolved with a PCR positivity curve *p*(*s*), the probability of someone infected at time *s* = 0 to test PCR positive to yield prevalence of PCR positivity *P* (*t*).

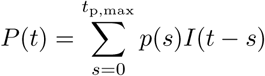

where *t*_p,max_ = 60 is the maximum time modelled for which a person can stay PCR positive. We assumed each *p*(*s*) to have an independent normal prior distribution at each time *s* after infection with given mean and standard deviation estimates from the posterior estimates of another study (Hellewell et al., 2021). Infection incidence *I*(*t*) is distinct from the estimates of PCR positivity incidence provided by ONS alongside the prevalence estimates, as it allows for the probability of infections yielding negative PCR results as a function of the time since infection and is indexed by date of infection rather than the date of first testing positive.

We used Gaussian Process (GP) priors to ensure smoothness of the estimates and deal with data gaps, whereby alternatively either *I*(*t*) is has a GP prior with exponential quadratic kernel. To reduce the computational requirements of our approach we used an approximate rather than exact GP (Riutort-Mayol et al., 2020).

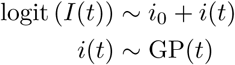

where *i*_0_ is the estimated mean of the GP, or the GP prior is applied to higher order differences when infections are non-stationary, for example the growth rate such as

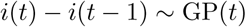

which implies that growth tends to zero when outside the range of the data, usually leading to better realtime performance (Abbott et al., 2020). The results shown here were obtained using this formulation with a GP prior on the growth rate.

We assumed that the probability of observing prevalence *Y*_P,*t*_ at time *t* was given by independent normal distributions with mean *P* (*t*) and standard deviation

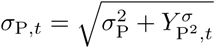

where *σ*_P_ was estimated as part of the inference procedure and 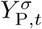 calculated based on the reported credible intervals in the ONS data, assuming independent normal errors. For data sets where only weekly estimates were reported by ONS, for example at the sub-regional level, we calculated average prevalence across the time period reported from our daily prevalence estimates.

Using the estimate infection incidences *I*(*t*) we estimated growth rates *r*(*t*) as

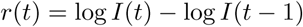

and reproduction numbers *R*(*t*) using the renewal equation as

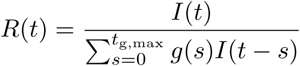

where *g*(*s*) is the distribution of the generation interval since the time of infection (Fraser, 2007). We assumed a maximum generation interval of *t*_g,max_ = 14. We use re-estimated generation intervals from early in the pandemic in Singapore as reported previously (Abbott et al., 2020).

#### Antibodies

When additionally using antibodies we convolve the modelled infections *I*(*t*) as well as input data on vaccinations *Y*_V,*t*_ with distributions quantifying the delay to generating detectable antibodies following infection (by default set to 4 weeks for both infection and vaccination), yielding potentially antibody-generating time series from infection *I*_A_ and *V*_A_. We then calculate antibodies from infection as

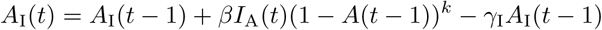

and antibodies from vaccination as

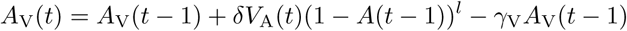

with the total population proportion with antibodies given as the sum of the two,

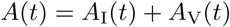

Here, the additional parameter *β* can be interpreted as proportion of new infections that does not increase the population proportion with antibodies, either due to lack of seroconversion or because they are breakthrough infections in those with existing antibodies, and parameters *k* and *l* govern the degree to which new seropositives preferentially arise in those not seropositive so far. Additional parameters *γ*_I_ and *γ*_V_ can be interpreted as rates of waning from natural infection and vaccination, respectively. This formulation implies simplifying assumptions that the rate of waning of detectable antibodies is exponential, that vaccine doses are allocated randomly amongst those with or without existing antibodies, and that the proportion of new vaccinations that lead to seroconversion *δ* is constant and independent of age, vaccine use, and dose number.

### Implementation

The model was implemented in *Stan* and using the *cmdstanr* R package (Gabry & Češnovar, 2021; Stan Development Team, 2022). All code needed to reproduce the results shown here is available at https://github.com/epiforecasts/inc2prev.

## Results

The model was able to reproduce the daily prevalence estimates and weekly antibody prevalence estimates published by ONS with reasonable accuracy when run until 15 November 2021 (Figure 1). The peaks of the corresponding incidence curve are earlier, higher and sharper. Estimated reproduction numbers highlight some key phases of the UK pandemic between April 2020 and November 2021, in particular rapid increases due to emergence of the Alpha variant in December followed by a period of low transmission during lockdown until March 2021, and rapid spread of the Delta variant in May-July 2021 followed by a period of relatively steady transmission.

**Figure 1:**
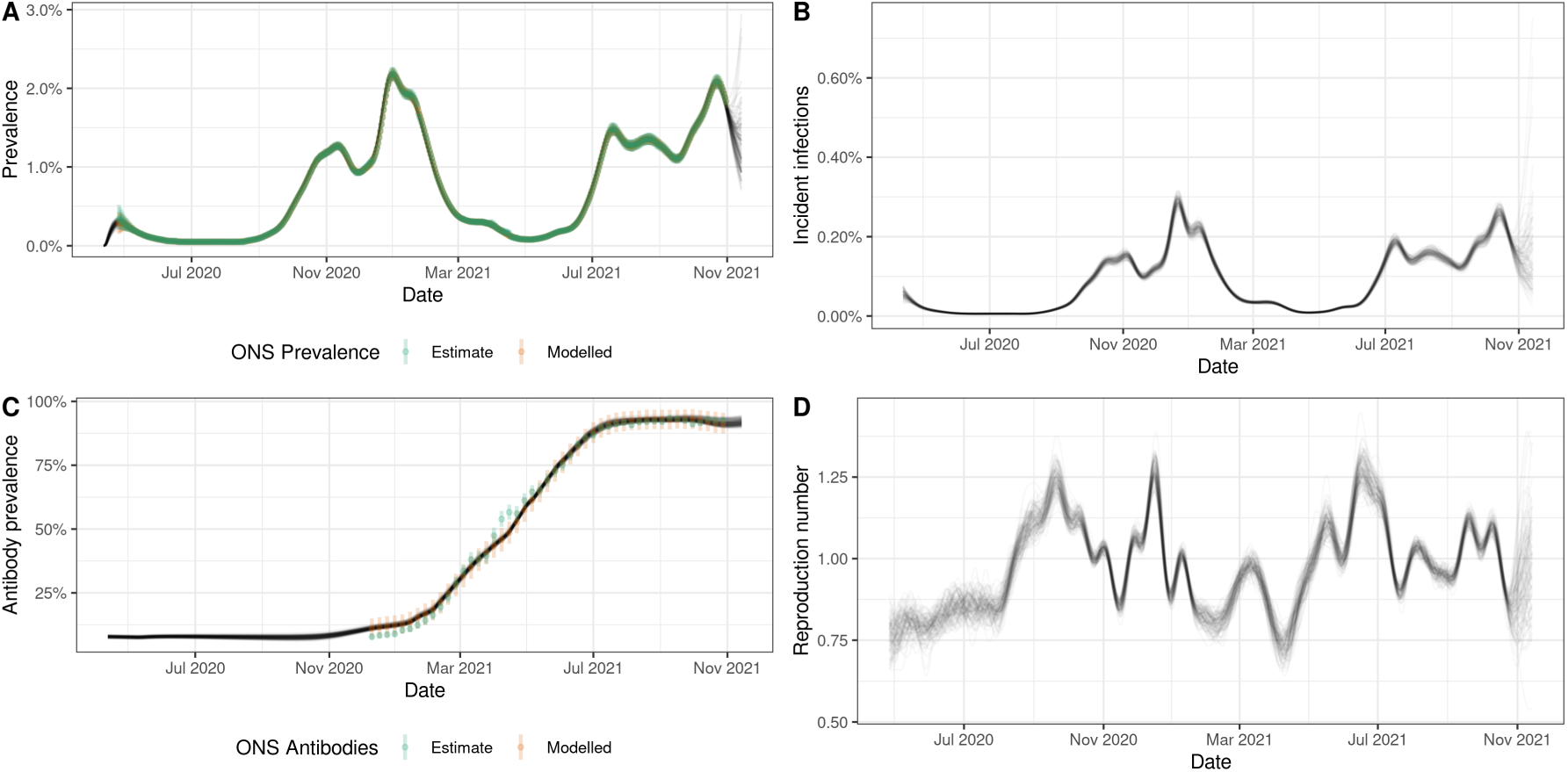
Model posteriors for England. A. Estimates of daily modelled prevalence and modelled prevalence as published by ONS. B. Estimated incidence of new infections. C. Estimated antibody prevalence and estimes as published by ONS. D. Estimated reproduction numbers.

Posterior estimates of recovered biological parameters are shown in Table 1. Some of the parameter estimates show high levels of correlation suggesting issues of identifiability (Figure 2).

**Table 1:** Estimates and credible intervals (CIs, as quantiles of the posterior distribution) of biological parameters.

| Parameter | Description | Estimate (90% CI) |
| --- | --- | --- |
| beta | Proportion infected that seroconvert | 0.82 (0.66–0.94) |
| delta | Proportion vaccinated that seroconvert | 0.98 (0.96–0.99) |
| gamma (infection) | Antibody waning following infection (per month) | 0.032 (0.0055–0.059) |
| gamma (vaccination) | Antibody waning following vaccination (per month) | 0.014 (0.0024–0.029) |
| k | Efficacy adjustment of immunity following infection | 1 (0.85–1.2) |
| l | Efficacy adjustment of immunity following vaccination | 0.67 (0.61–0.75) |

**Figure 2:**
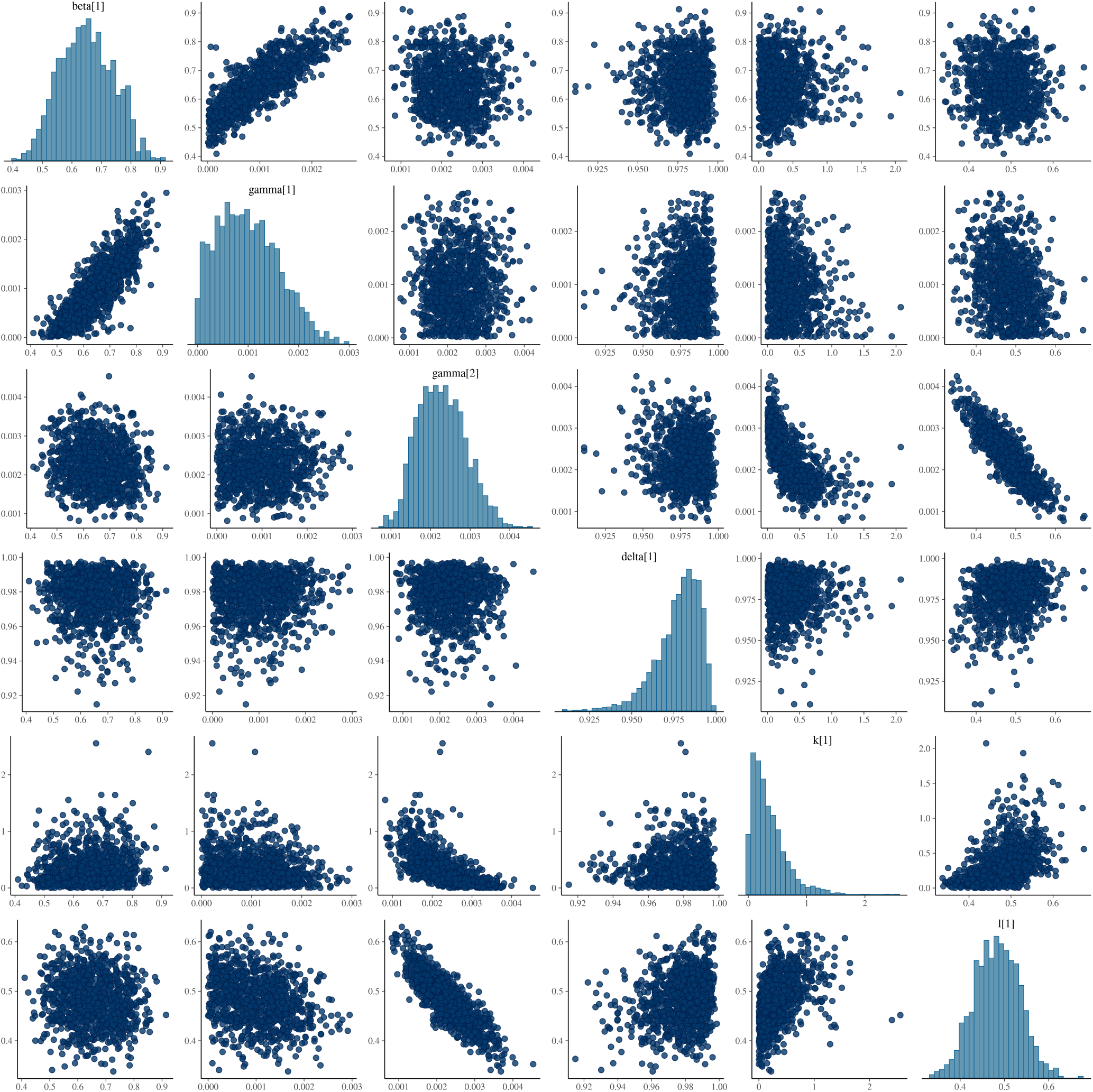
Scatter plots and histograms of posterior parameter samples.

## Discussion

We have presented a method to estimate epidemiological parameters such as infection incidence, timevarying reproduction numbers and growth rates from repeated cross-sectional PCR positivity estimates. The estimates of infection incidence are distinct from estimates of PCR positivity incidence that are reported alongside the positivity prevalence estimates, as the probability of detecting infections is low early in the course of an infection, and more generally varies over said course (Hellewell et al., 2021). When additionally using antibody and vaccination data, we refine our estimates of infection incidence and recover estimates of relevant parameters such as seroconversion and waning rates that can be used to estimate antibody prevalence where infection and vaccination data is available but antibody data is not.

Our estimated parameters of antibody dynamics are averages across a various combinations of vaccine types and individual factors that are known to affect immunological responses to either infection or vaccination, particularly age (Ward et al., 2022). We estimated that 18% (90% credible interval, CI: 6–34) of individuals do not seroconvert after infection, consistent with the 24% estimated from the same study population, but also lower estimates such as 10% in a different study (Gudbjartsson et al., 2020). We further estimated that 98% (90% CI: 99–96) of individuals seroconverted following vaccination, in line with high such proportions estimated in healthcare workers (Eyre et al., 2021). Our estimates of waning suggest that detectable antibodies decrease by 3.2% (90% CI: 0.55–5.9) following infection and 1.4% (90% CI: 0.24–2.9) following vaccination. All these values depend on the specific cutoff used for seropositivity and combine a range of vaccines, and they ignore additional effects from receiving multiple doses of vaccine, becoming infected as well as vaccinated. They cannot be compared directly to estimates of vaccine efficacy or waning thereof.

As currently implemented, our method suffers from a number of limitations that risk biasing the results. Several of the key parameters in our model, especially the estimates of PCR positivity over time from infection, generation interval distributions, are fixed and based on estimates derived from wildtype virus in a particular cohort of healthcare workers and may well be incorrect for other circulating variants or populations. Furthermore, generation times have been shown to change over time due to behavioural changes and epidemiological dynamics, which would affect our reproduction number estimates (Champredon & Dushoff, 2015; Hart et al., 2021; Park et al., 2021). PCR detection probabilities as a function of time since infection were based on independent normal distributions, whereas in reality they are likely to be correlated over time. We modelled the growth of infections as a stationary Gaussian process, whereas in reality variation over time has changed between periods of stability and rapid change due to changes in contact behaviour in response to the epidemic. Lastly, we assumed that antibody waning was exponential, and ignored any consequences of multiple rounds of vaccination or infection apart from converting those without detectable antibodies to having detectable antibodies.

Future directions of this work should help address some of these limitations, for example by including more detail on antibody levels, or by including antibody measurements that may be able to distinguish between natural and vaccine-acquired immunity (Amjadi et al., 2021). It could further make use of more comprehensive information on PCR detection curves taking into account correlations in detectability since time from infection and pointly jointly estimating these curves using individual level data. Combined with other data streams, for example on test-positive community cases, or severe outcomes resulting in hospitalisations or deaths, our method could be used to understand rates of notification or sever disease given infection, or to generate forecasts of expected burden. Lastly, more detailed information on the infections detected, for example viral loads via Cycle threshold (Ct) values, could be used to improve real-time performance of growth rates and reproduction numbers (Hay et al., 2021).

There is enormous potential for understanding epidemiological dynamics from repeated cross-sectional surveys, whether to identify current or past infection (Metcalf et al., 2016). Where the generation interval distribution is the same or close to the distribution of detectability after infection, this could be done using recently developed methods for unified modelling of incidence and prevalence (Pakkanen et al., 2021). The methods presented here and related ones could be applied to other infections monitored in a similar way, and thus in combination with such data collection and publication become a tool for monitoring epidemic and endemic infectious diseases in the future.

## Data Availability

All code needed to reproduce the results shown in the article are available at https://github.com/epiforecasts/inc2prev.

## Acknowledgements

We thank Thomas House for insightful comments on this work, and the Office for National Statistics for making the data sets publicly available. We acknowledge funding by the Wellcome Trust (210758/Z/18/Z).

## References

Abbott, S., Hellewell, J., Thompson, R. N., Sherratt, K., Gibbs, H. P., Bosse, N. I., Munday, J. D., Meakin, S., Doughty, E. L., Chun, J. Y., Chan, Y.-W. D., Finger, F., Campbell, P., Endo, A., Pearson, C. A. B., Gimma, A., Russell, T., Flasche, S., Kucharski, A. J., … Funk, S. (2020). Estimating the time-varying reproduction number of sars-cov-2 using national and subnational case counts. Wellcome Open Research, 5, 112. https://doi.org/10.12688/wellcomeopenres.16006.2

Amjadi, M. F., Adyniec, R. R., Gupta, S., Bashar, S. J., Mergaert, A. M., Braun, K. M., Moreno, G. K., O’Connor, D. H., Friedrich, T. C., Safdar, N., McCoy, S. S., & Shelef, M. A. (2021). Anti-membrane and anti-spike antibodies are long-lasting and together discriminate between past covid-19 infection and vaccination. https://doi.org/10.1101/2021.11.02.21265750

Champredon, D., & Dushoff, J. (2015). Intrinsic and realized generation intervals in infectious-disease transmission. Proceedings of the Royal Society B: Biological Sciences, 282 (1821), 20152026. https://doi.org/10.1098/rspb.2015.2026

Eyre, D. W., Lumley, S. F., Wei, J., Cox, S., James, T., Justice, A., Jesuthasan, G., O’Donnell, D., Howarth, A., Hatch, S. B., Marsden, B. D., Jones, E. Y., Stuart, D. I., Ebner, D., Hoosdally, S., Crook, D. W., Peto, T. E., Walker, T. M., Stoesser, N. E., … Jeffery, K. (2021). Quantitative sars-cov-2 anti-spike responses to pfizer–biontech and oxford–astrazeneca vaccines by previous infection status. Clinical Microbiology and Infection, 27 (10), 1516.e7–1516.e14. https://doi.org/10.1016/j.cmi.2021.05.041

Fraser, C. (2007). Estimating individual and household reproduction numbers in an emerging epidemic. PLoS ONE, 2 (8), e758. https://doi.org/10.1371/journal.pone.0000758

Gabry, J., & Češnovar, R. (2021). Cmdstanr: R interface to ‘cmdstan’.

Gudbjartsson, D. F., Norddahl, G. L., Melsted, P., Gunnarsdottir, K., Holm, H., Eythorsson, E., Arnthorsson, A. O., Helgason, D., Bjarnadottir, K., Ingvarsson, R. F., Thorsteinsdottir, B., Kristjansdottir, S., Birgisdottir, K., Kristinsdottir, A. M., Sigurdsson, M. I., Arnadottir, G. A., Ivarsdottir, E. V., Andresdottir, M., Jonsson, F., … Stefansson, K. (2020). Humoral immune response to sars-cov-2 in iceland. New England Journal of Medicine, 383 (18), 1724–1734. https://doi.org/10.1056/nejmoa2026116

Hart, W. S., Abbott, S., Endo, A., Hellewell, J., Miller, E., Andrews, N., Maini, P. K., & Thompson, R. N. (2021). Inference of sars-cov-2 generation times using uk household data. https://doi.org/10.1101/2021.05.27.21257936

Hay, J. A., Kennedy-Shaffer, L., Kanjilal, S., Lennon, N. J., Gabriel, S. B., Lipsitch, M., & Mina, M. J. (2021). Estimating epidemiologic dynamics from cross-sectional viral load distributions. Science, 373 (6552). https://doi.org/10.1126/science.abh0635

Hellewell, J., Russell, T. W., Beale, R., Kelly, G., Houlihan, C., Nastouli, E., & Kucharski, A. J. (2021). Estimating the effectiveness of routine asymptomatic pcr testing at different frequencies for the detection of sars-cov-2 infections. BMC Medicine, 19 (1). https://doi.org/10.1186/s12916-021-01982-x

Metcalf, C. J. E., Farrar, J., Cutts, F. T., Basta, N. E., Graham, A. L., Lessler, J., Ferguson, N. M., Burke, D. S., & Grenfell, B. T. (2016). Use of serological surveys to generate key insights into the changing global landscape of infectious disease. The Lancet, 388 (10045), 728–730. https://doi.org/10.1016/s0140-6736(16)30164-7

Murray, J., & Cohen, A. L. (2017). Infectious disease surveillance. International Encyclopedia of Public Health, 222–229. https://doi.org/10.1016/b978-0-12-803678-5.00517-8

Pakkanen, M. S., Miscouridou, X., Berah, T., Mishra, S., Mellan, T. A., & Bhatt, S. (2021). Unifying incidence and prevalence under a time-varying general branching process. http://arxiv.org/abs/2107.05579v2

Park, S. W., Bolker, B. M., Funk, S., Metcalf, C. J. E., Weitz, J. S., Grenfell, B. T., & Dushoff, J. (2021). Roles of generation-interval distributions in shaping relative epidemic strength, speed, and control of new sars-cov-2 variants. https://doi.org/10.1101/2021.05.03.21256545

Pouwels, K. B., House, T., Robotham, J. V., Birrell, P. J., Gelman, A., Bowers, N., Boreham, I., Thomas, H., Lewis, J., Bell, I., Bell, J. I., Newton, J. N., Farrar, J., Diamond, I., Benton, P., & Walker, A. S. (2020). Community prevalence of sars-cov-2 in england: Results from the ons coronavirus infection survey pilot. https://doi.org/10.1101/2020.07.06.20147348

Riutort-Mayol, G., Bürkner, P.-C., Andersen, M. R., Solin, A., & Vehtari, A. (2020). Practical hilbert space approximate bayesian gaussian processes for probabilistic programming. http://arxiv.org/abs/2004.11408

Stan Development Team. (2022). Stan Modeling Language Users Guide and Reference Manual, Version 2.29. http://mc-stan.org/

Ward, H., Whitaker, M., Flower, B., Tang, S. N., Atchison, C., Darzi, A., Donnelly, C. A., Cann, A., Diggle, P. J., Ashby, D., Riley, S., Barclay, W. S., Elliott, P., & Cooke, G. S. (2022). Population antibody responses following covid-19 vaccination in 212,102 individuals. Nature Communications, 13 (1). https://doi.org/10.1038/s41467-022-28527-x

